# Electrocardiography-Based Prediction of Sudden Cardiac Death in Heart Failure Patients: Application of Artificial Intelligence

**DOI:** 10.1101/2022.03.20.22272659

**Authors:** Yasuyuki Shiraishi, Shinichi Goto, Nozomi Niimi, Yoshinori Katsumata, Ayumi Goda, Makoto Takei, Mike Saji, Yosuke Nishihata, Motoaki Sano, Keiichi Fukuda, Takashi Kohno, Tsutomu Yoshikawa, Shun Kohsaka

**Affiliations:** Department of Cardiology, Keio University School of Medicine, Tokyo, Japan; Institute for Integrated Sports Medicine, Keio University School of Medicine, Tokyo, Japan; One Brave Idea and Division of Cardiovascular Medicine, Department of Medicine, Brigham and Women’s Hospital, Boston, MA, USA; Department of Cardiovascular Medicine, Kyorin University School of Medicine, Tokyo, Japan; Department of Cardiology, Saiseikai Central Hospital, Tokyo, Japan; Department of Cardiology, Sakakibara Heart Institute, Tokyo, Japan; Department of Cardiology, St. Luke’s International Hospital, Tokyo, Japan

**Keywords:** artificial intelligence, electrocardiogram, heart failure, left ventricular ejection fraction, sudden cardiac death

## Abstract

**Background:** Although predicting sudden cardiac death (SCD) in patients with heart failure (HF) is critical, the current predictive model is suboptimal. Electrocardiography-based artificial intelligence (ECG-AI) algorithms may better stratify risk. We assessed whether the ECG-AI index established here could better predict SCD in HF and whether the ECG-AI index and conventional predictors of SCD can improve SCD stratification.

**Methods:** In a prospective observational study, four tertiary care hospitals in metropolitan Tokyo that enrolled 2,559 patients hospitalized with HF who were successfully discharged after acute decompensation. ECG data collected during the index hospitalization were extracted from the hospitals’ electronic medical record systems. The ECG-AI index is the output from an AI model that was trained to predict the risk of SCD based on ECG input. The association between ECG-AI index and SCD was evaluated with adjustment for left ventricular ejection fraction (LVEF), New York Heart Association (NYHA) class, and competing risk of non-SCD. The outcome measure was a composite of SCD and implantable cardioverter-defibrillator activation. The ECG-AI index was established using a derivation (hospital A) and validation cohort (hospital B), and its ability was evaluated in a test cohort (hospitals C and D).

**Results:** The ECG-AI index plus classical predictive guidelines (i.e., LVEF ≤ 35%, NYHA class II–III) significantly improved the discriminative value of SCD (area under the receiver operating characteristic curve, 0.66 vs. 0.59; p=0.017; Delong’s test) with good calibration (p=0.11; Hosmer–Lemeshow test) and improved net reclassification (36%; 95% confidence interval, 9%–64%; p=0.009). The Fine-Gray model considering the competing risk of non-SCD demonstrated that the ECG-AI index was independently associated with SCD (adjusted sub-distributional hazard ratio, 1.25; 95% confidence interval, 1.04–1.49; p=0.015). An increased proportional risk of SCD vs. non-SCD with increasing ECG-AI index was also observed (low, 16.7%; intermediate, 18.5%; high, 28.7% risk; p for trend = 0.023). Similar findings were observed in patients aged ≤75 years with a non-ischemic etiology and an LVEF >35%.

**Conclusions:** Among patients with HF, ECG-based AI significantly improved the SCD risk stratification compared to the conventional indication for implantable cardioverter-defibrillators inclusive of LVEF and NYHA class.

## INTRODUCTION

Heart failure (HF) is a serious clinical condition associated with poor patient quality of life and premature death; approximately 50% of patients with HF die within 5 years of diagnosis.^1,2^ A high incidence of HF is observed in Asia (1.2%–6.7%) and Western countries (1%–14%),^2-5^ and as the prevalence is expected to increase, HF creates a substantial global public health burden. Sudden cardiac death (SCD), typically caused by lethal arrhythmias, is reportedly responsible for approximately 50% of all cardiovascular deaths in patients with HF,^6,7^ and contributes substantially to this health burden. Although implantable cardioverter-defibrillators (ICDs) are used to reduce the risk of SCD, the implantation procedure is invasive, and approximately 50% of patients with ICD implantation experience inappropriate shocks, reducing quality of life and increasing mortality rates.^8,9^ Therefore, the accurate assessment of SCD risk in patients with HF is paramount for clinical decision-making to ensure appropriate device application.

Current approaches to assessing SCD risk are mainly based on left ventricular ejection fraction (LVEF; ≤ 35%) and the New York Heart Association (NYHA) functional classification and remain suboptimal,^10,11^ resulting in an over- and underuse of ICD.^12^ Furthermore, despite the fact that SCD also occurs frequently in patients without severe left ventricular dysfunction, there is currently no risk stratification algorithm for SCD in patients with an LVEF > 35%.^13^ Artificial intelligence (AI) is a promising technology for deriving a statistical model from information-rich yet complex datasets.^14^ Electrocardiography-based AI (ECG-AI) models constructed by convolutional neural networks (CNNs) have shown potential to detect disease,^15^ predict cardiac function,^16-18^ and estimate prognosis.^19^

Therefore, we hypothesized that AI models trained on ECG will enable the detection of important features for classifying the risk of SCD and improve the risk stratification of patients with HF, allowing for a more appropriate application of medical resources. Using data from a prospective observational study, we aimed to assess the ability of ECG-based AI to predict the incidence of SCD among patients with HF who were hospitalized for acute decompensation, required urgent treatment, and were successfully discharged.

## METHODS

### Data Source and Data Collection

The design of the West Tokyo Heart Failure Registry (WET-HF) has been described previously (eMethod 1 in the **Supplement**).^20,21^ Using data from this prospective observational study, we analyzed hospitalized patients with HF who required urgent treatment for acute decompensation in four tertiary care hospitals (Keio University Hospital, Kyorin University Hospital, Sakakibara Heart Institute, and St. Luke’s International Hospital) within the metropolitan area of Tokyo, Japan. All 12-lead ECG data of the patients were reported and extracted from the electronic medical record system of each institution. A standard 12-lead ECG has 15 voltage-time traces, including those 2.5 s in duration for all 12 leads and those 10 s in duration for leads V1, II, and V5. The ECG data were stored as measurements of the time-series voltage at a sampling rate of 500 Hz. Of the six hospitals, two were not included in the present study because the ECG data could not be extracted from the database. Patients who were registered in the remaining hospitals underwent ECG at the time of discharge.

### Study Cohort

The patient selection and exclusion criteria as well as the group allocations are shown in eFigure 1 in the **Supplement**. A total of 2,559 patients who were successfully discharged between 2006 and 2017 were included in the present analysis. Patients were allocated to one of the three cohorts (derivation, validation, or test) based on the hospital of recruitment. To maximize the data for model training, the derivation cohort consisted of patients from two institutions, while the validation and test cohorts consisted of patients from only one institution. The ECG data of the patients at the time of discharge were reported and extracted from the electronic medical record systems. The latest data were included in the analysis when multiple ECG data were available for the same patient. By design, only one ECG reading was used for each patient; thus, no data from a single patient were allocated to more than two cohorts.

**Figure 1.**
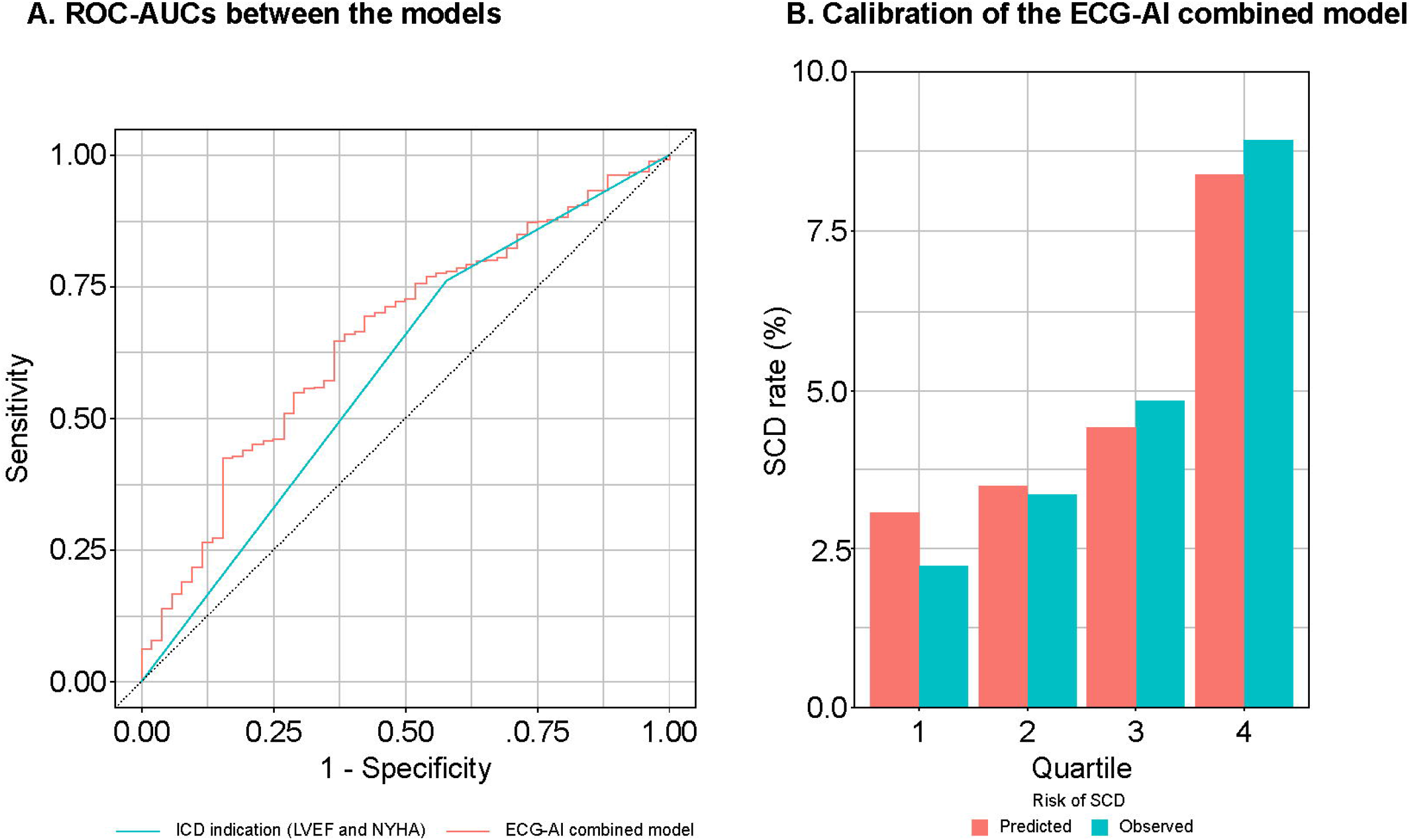
Discrimination and calibration of risk models for predicting SCD events. Comparison of ROC-AUC (a) shows the conventional guideline–directed indication for ICD (LVEF ≤ 35% and NYHA class II–III; AUC = 0.59 [95% CI, 0.52–0.66]) vs. the ECG-AI combined model (ECG-AI index + ICD indication; AUC = 0.66 [95% CI, 0.58–0.73]) for predicting 3-year composite SCD events (p = 0.017 for Delong’s test). Calibration of the ECG-AI combined model is shown (b) by dividing four bins based on the quartiles of the model-predicted risk (p = 0.11 for the Hosmer–Lemeshow test). CI, confidence interval; ECG-AI, electrocardiogram-based artificial intelligence; ICD, implantable cardioverter-defibrillator; LVEF, left ventricular ejection fraction; NYHA, New York Heart Association; ROC-AUC, receiver operating characteristic area under the curve; SCD, sudden cardiac death

### Model Training

The AI model to predict 3-year SCD was constructed using a combination of CNN and long short-term memory (LSTM), a variant of recurrent neural network (RNN) (eFigure 2 in the **Supplement**). The details of the architecture have been published previously.^22^ The architecture consists of a neural network that stacks up four layers of one-dimensional CNN suitable for detecting “shape patterns” with a relatively low computational cost and two layers of LSTM suitable for learning time-series data in detail with a relatively high computational cost. The model was trained with the data from the derivation cohort to minimize the binary cross-entropy loss with the RMSProp optimizer with an initial learning rate of 0.0001. The performance of the model was calculated using data from the validation dataset at the end of each epoch. The final model was chosen as the one that performed best for 50 epochs in the validation cohort (eFigure 3 in the **Supplement**). To ensure that the model works on data that were never seen during training and the model selection procedure, the performance of the final model (i.e., the ECG-AI index) was calculated only once using data from the test cohort. Finally, we implemented gradient-weighted class activation mapping (Grad-CAM) to identify which regions in the ECG were based on the prediction of the neural network model (eFigure 4 in the **Supplement**). The model was trained using Tensorflow framework version 2.2.0, with Python version 3.6.8.

**Figure 2.**
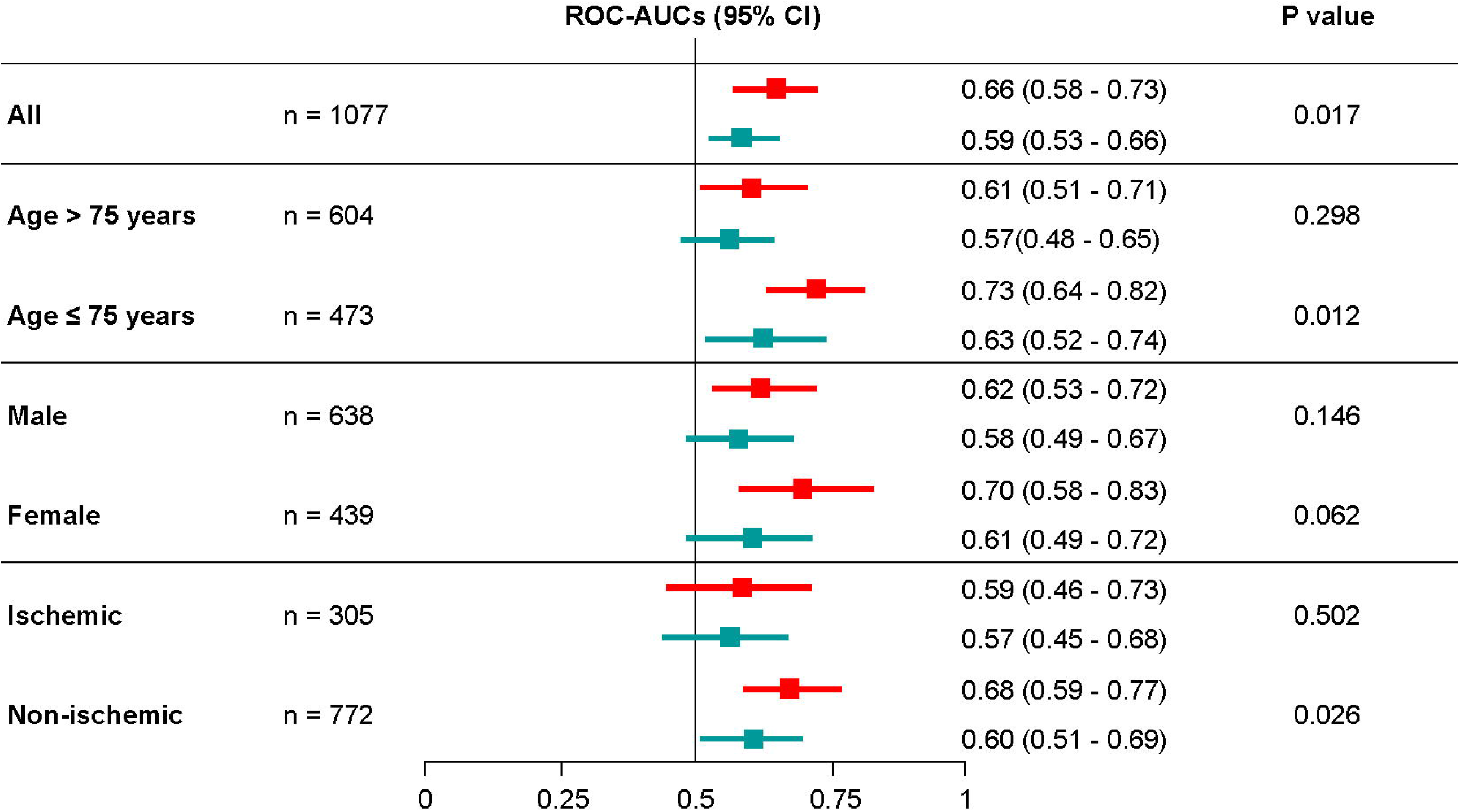
Subgroup analyses of the discriminative abilities of the ECG-AI combined model and conventional indication for ICD. The model’s discrimination was compared to that of the Delong test. Red bars, the ECG-AI combined model; Green bars, the conventional indication for ICD. CI, confidence interval; ECG-AI, electrocardiogram-based artificial intelligence; ICD, implantable cardioverter-defibrillator; ROC-AUC, receiver operating characteristic area under the curve

**Figure 3.**
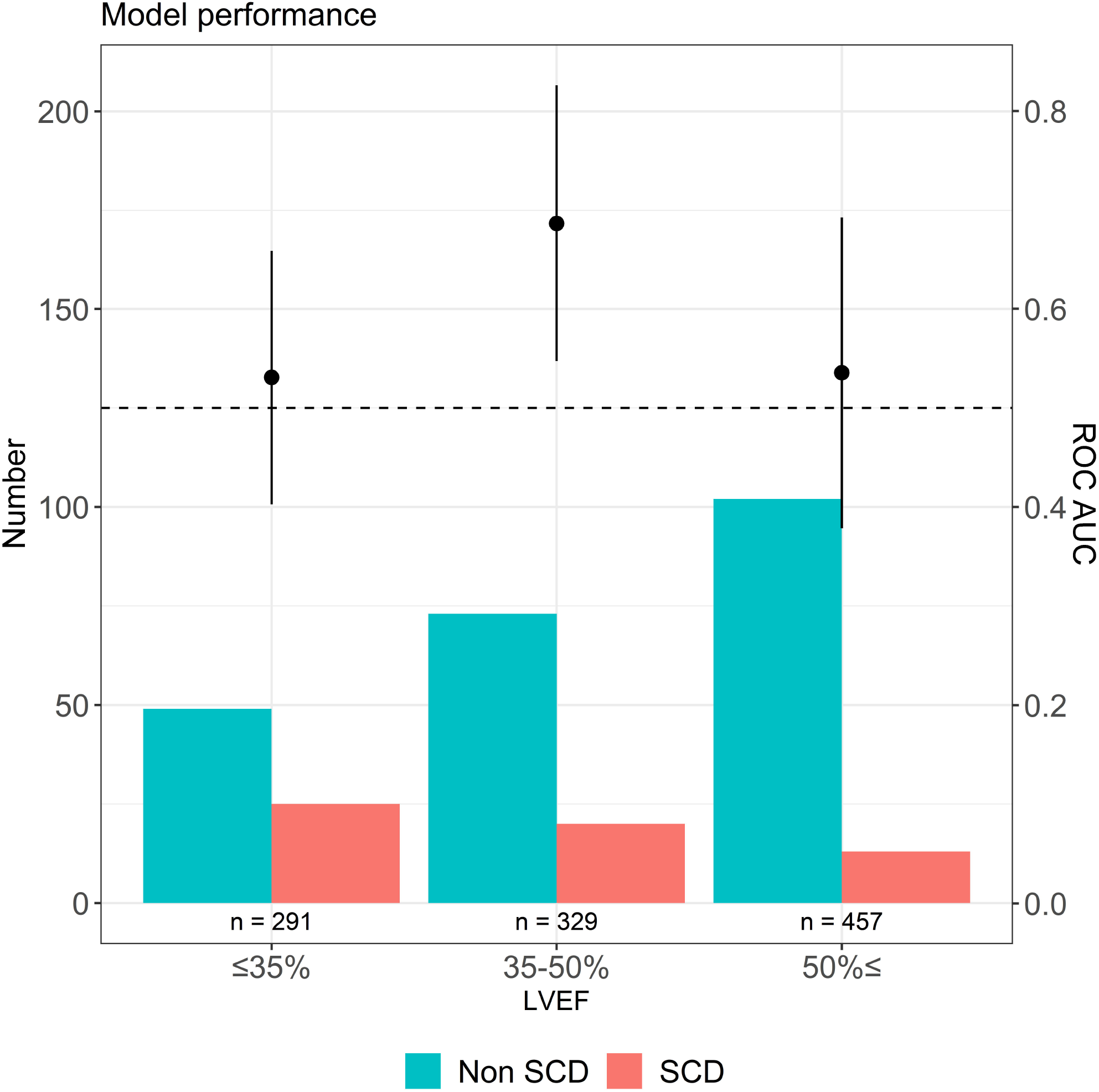
Discriminative ability of the ECG-AI index and frequency of SCD vs. non-SCD events by LVEF category. The above forest plots show ROC-AUC with 95% confidence intervals of the ECG-AI index for predicting SCD events by LVEF. In patients with an LVEF of 35%–50%, the ECG-AI index showed the best discriminative ability. The bar graphs represent the number of SCD and non-SCD patients according to LVEF. The proportion of patients with SCD vs. non-SCD steadily decreased as LVEF increased. ECG-AI, electrocardiogram-based artificial intelligence; LVEF, left ventricular ejection fraction; ROC-AUC, receiver operating characteristic area under the curve; SCD, sudden cardiac death

**Figure 4.**
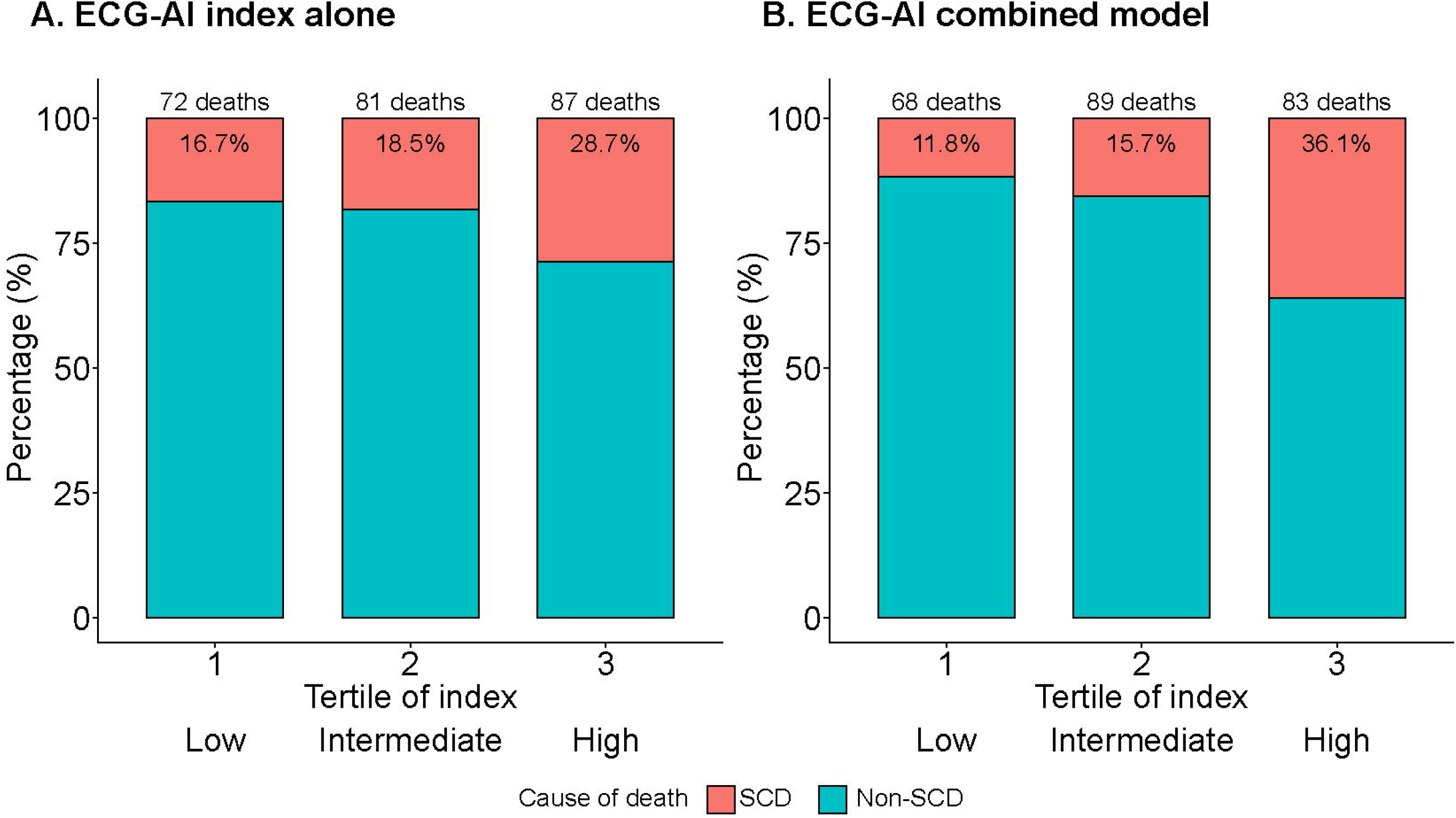
Proportional risk of SCD vs. non-SCD events based on the model-based risk. ECG-AI index (a), ECG-AI combined model (with LVEF and NYHA class b). The patients were divided into three groups by tertile of risk score (each p for trend < 0.05). ECG-AI, electrocardiogram-based artificial intelligence; LVEF, left ventricular ejection fraction; NYHA, New York Heart Association; SCD, sudden cardiac death. -AUC, receiver operating characteristic area under the curve

### Ascertainment and Classification of SCD or ICD Events

The outcome measure was a composite of SCD and ICD activation (i.e., both shock and anti-tachycardia pacing). To ensure the accuracy of SCD assessment, the WET-HF registry was supported by a central study committee that adjudicated the mode of death. All deaths were reviewed by the investigators and then categorized into those in need of adjudication or those in which the mode of death could be defined clearly. Central committee members reviewed the abstracted records and adjudicated modes of death. SCD was defined as unexpected and otherwise unexplained death in a previously stable patient or death from documented or presumed cardiac arrhythmia without a clear non-cardiovascular cause, including patients who were comatose and then died after attempted resuscitation.^23^ Patients who died and had been out of contact for more than 24 h were classified as “unknown death.”^23^ All other causes of death were classified as non-SCD. In addition, ICD activation was ascertained through device interrogation during regular check-ups or at a suspected instance of an arrhythmic episode and SCD.

### Statistical Analyses

With respect to descriptive statistics, continuous variables are presented as median and interquartile range, while categorical variables are presented as frequency and percentage. For baseline characteristics, the three cohorts (i.e., derivation, validation, and test) were compared using the Kruskal–Wallis rank sum test for continuous variables and the chi-square test or Fisher’s exact test for categorical variables as appropriate.

The discriminative ability of the conventional guideline–directed indication for ICD (i.e., LVEF ≤ 35% and NYHA class II–III) or its combination with the ECG-AI index (ECG-AI combined model) for predicting composite SCD events over 3 years was evaluated using the receiver operating characteristic area under the curve (ROC-AUC) with logistic regression analysis and the pairwise Delong’s test. The model’s calibration performance was assessed by comparing the predicted and observed probabilities for the four groups using the Hosmer–Lemeshow test. The model’s reclassification of the composite SCD events was also assessed as a net reclassification improvement using the 3-year estimated probabilities of composite SCD events.

Next, the model’s discriminative abilities were assessed in the pre-specified subgroups (i.e., age (≥ 75 vs. < 75 years), sex (male vs. female), and etiology (ischemic vs. non-ischemic). In the subset of patients divided by LVEF, the discriminative ability of the ECG-AI index, as well as the frequencies of composite SCD vs. non-SCD events were assessed separately: ≤ 35%, 35%–50%, and ≥ 50%. Furthermore, we performed sensitivity analyses using several LVEF cut-off values (45%, 55%, and 60%).

For the survival analysis, we evaluated the cumulative incidence of composite SCD and non-SCD events using the Aalen–Johansen estimator divided by the risk of the conventional guideline–directed indication for ICD and the ECG-AI combined model. The optimal thresholds for risk categories (low vs. high risk) for ICD indication and the ECG-AI combined model were determined using the Youden index. We also examined the association between the ECG-AI index and composite SCD events using univariate and multivariate Fine and Gray models, which accounted for the competing risk of non-SCD. To assess the model’s discrimination of the survival analysis, we used Harrel’s c-statistics for the Fine and Gray model. We transformed the ECG-AI index (the output of the neural network model) to be standardized (mean = 0; standardized deviation = 1). A survival analysis that did not consider the competing risk was also performed with time-dependent ROC-AUC.

To visualize the proportional risk of composite SCD vs. non-SCD events, we divided the patients into three groups according to tertile of predicted risk by the ECG-AI index alone and the ECG-AI combined model and calculated the prevalence of SCD and non-SCD events by group using a trend test. All analyses were conducted using the tidyverse, tidymodels, pROC, PredictABEL, survminer, cmprsk, riskRegression, and survival packages of R version 4.0.3 (R Foundation for Statistical Computing, Vienna, Austria, 2008).

## RESULTS

### Patient Characteristics

The baseline characteristics of the derivation, validation, and test cohorts are shown in **Table 1**. The mean patient age and LVEF were 73–78 years and 40%–48%, respectively. The proportions of patients with NYHA functional class II–III were 75.2%, 92.4%, and 93.4% in the derivation, validation, and test cohorts, respectively. Medical therapies for HF were similarly implemented during the index hospitalization across each cohort: angiotensin-converting enzyme inhibitors or angiotensin receptor blockers, 62.3%–69.2%; beta blockers, 75.7%–79.2%; and mineralocorticoid receptor antagonists, 29.9%–34.9%. The prevalence of ICD implantation was low at 3.0%–8.4%. Overall, 236 (21.9%) deaths (48 [20.3%] SCDs and 188 [79.7%] non-SCDs) and 4 ICD activations occurred in 1,077 patients who were included in the test cohort in the 3 years following the hospitalization events.

**Table 1.** Patient backgrounds according to derivation, validation, and test cohorts.

| Value | Derivation cohort<br>n = 1,028 | Validation cohort<br>n = 454 | Test cohort<br>n = 1,077 | P-value |
| --- | --- | --- | --- | --- |
| Age, years | 78 (69–84) | 73 (60–81) | 77 (67–84) | < 0.001 |
| Men, n (%) | 606 (58.9) | 288 (63.4) | 638 (59.2) | 0.400 |
| Body mass index, kg/m<sup>2</sup> | 21.5 (19.3–24.1) | 21.8 (19.5–24.1) | 21.1 (18.7–23.8) | < 0.001 |
| Systolic blood pressure, mm Hg | 110 (100–120) | 108 (97–120) | 113 (100–128) | < 0.001 |
| Heart rate, bpm | 70 (60–78) | 71 (61–80) | 70 (61–80) | < 0.001 |
| LVEF, % | 48 (32–59) | 40 (30–56) | 48 (34–59) | < 0.001 |
| LVEF ≤ 35%, n (%) | 321 (31.5) | 209 (46.2) | 290 (26.9) | < 0.001 |
| NYHA class II–III, n (%) | 771 (75.2) | 416 (92.4) | 990 (93.4) | < 0.001 |
| Previous HF hospitalization | 337 (32.7) | 146 (31.7) | 257 (27.5) | < 0.001 |
| Atrial fibrillation, n (%) | 607 (59.0) | 210 (46.1) | 428 (39.7) | < 0.001 |
| Hypertension, n (%) | 653 (63.5) | 266 (58.5) | 766 (71.1) | < 0.001 |
| Diabetes mellitus, n (%) | 296 (28.8) | 145 (31.8) | 428 (39.7) | < 0.001 |
| Stroke, n (%) | 140 (12.9) | 66 (14.2) | 141 (13.1) | 0.552 |
| COPD, n (%) | 27 (2.5) | 42 (7.4) | 67 (6.0) | < 0.001 |
| Hemoglobin, g/dL | 11.9 (10.5–13.3) | 12.5 (10.7–14.3) | 11.9 (10.6–13.4) | < 0.001 |
| Blood urea nitrogen, mg/L | 20.1 (15.2–29.4) | 23.9 (18.0–32.2) | 24.4 (17.9–35.2) | < 0.001 |
| eGFR, ml/min/1.73 m<sup>2</sup> | 51.8 (37.7–64.5) | 50.3 (36.3–62.4) | 49.2 (30.9–66.0) | 0.001 |
| Sodium, mEq/L | 139 (137–141) | 139 (137–141) | 139 (136–141) | 0.522 |
| Potassium, mEq/L | 4.3 (4.0–4.6) | 4.4 (4.1–4.7) | 4.3 (4.0–4.7) | < 0.001 |
| Uric acid, mg/L | 6.6 (5.3–7.8) | 6.9 (5.6–8.2) | 7.2 (5.7–8.6) | < 0.001 |
| Albumin, mg/L | 3.6 (3.3–3.9) | 3.7 (3.3–4.0) | 3.4 (3.0–3.7) | < 0.001 |
| BNP, pg/mL* | N/A | 244 (116–479) | 247 (128–508) |  |
| NT-proBNP, pg/mL* | 1,830 (1,109–3,612) | N/A | N/A |  |
| Loop diuretics, n (%) | 820 (79.8) | 315 (69.2) | 821 (76.2) | < 0.001 |
| ACEi or ARB, n (%) | 640 (62.3) | 315 (69.2) | 687 (63.8) | 0.018 |
| Beta blocker, n (%) | 788 (76.7) | 360 (79.2) | 815 (75.7) | 0.304 |
| MRA, n (%) | 307 (29.9) | 193 (34.9) | 378 (34.9) | < 0.001 |
| Digitalis, n (%) | 73 (7.1) | 54 (7.0) | 77 (7.0) | 0.011 |
| Amiodarone, n (%) | 75 (7.2) | 42 (8.8) | 114 (10.5) | 0.044 |
| ICD, n (%) | 52 (5.1) | 38 (8.4) | 32 (3.0) | < 0.001 |

BNP was measured in the validation and test cohorts, while NT-proBNP was measured in the derivation cohort.
Abbreviations: LVEF: Left ventricular ejection fraction, NYHA: New York Heart Association, HF: Heart failure, COPD: Chronic obstructive pulmonary disease, eGFR: Estimated glomerular filtration rate, BNP: B-type natriuretic peptide, NT-proBNP: N-terminal pro-B-type natriuretic peptide, ACEi: Angiotensin-converting enzyme inhibitor, ARB: Angiotensin receptor blocker, MRA: Mineralocorticoid receptor antagonist, ICD: Implantable cardioverter-defibrillator.

### ECG-Based AI Performance

The ROC curve analysis, without considering time-to-event, showed a good discriminative ability for predicting SCD over 3 years of the ECG-AI index with 0.62 (95% confidence interval [CI], 0.54–0.70) of ROC-AUC compared to the conventional guideline–directed indication for ICD. **Figure 1A** shows that the addition of the ECG-AI index to the conventional indication for ICD, inclusive of LVEF and NYHA class (ROC-AUC = 0.59 [95% CI, 0.52–0.66]), significantly improved the discrimination (ROC-AUC = 0.66 [95% CI, 0.58–0.73], p = 0.017, Delong’s test) compared to the conventional indication for ICD alone. The ECG-AI combined model showed good calibration (**Figure 1B**; p = 0.11, Hosmer–Lemeshow test). Furthermore, the addition of the ECG-AI index to the conventional indication for ICD improved the indices of reclassification (net reclassification improvement, 36% [9%–64%; p = 0.009]).

Similar findings were observed in the pre-specified subgroup analyses (**Figure 2**). In particular, compared to the conventional indication for ICD, the ECG-AI combined model showed significantly better discrimination of the incidence of SCD among younger patients (≤ 75 years) with a non-ischemic etiology.

The proportion of patients with SCD vs. non-SCD steadily decreased as LVEF increased, and the ECG-AI index showed the best discriminative ability in patients with an LVEF of 35%–50% **(Figure 3**). In sensitivity analyses using several LVEF cut-offs, the ECG-AI index showed the best performance for patients with LVEF 35%–50%, although a good discriminative ability was also observed among those with an LVEF of 35%–60% (eTable 1 in the **Supplement**). In addition, another sensitivity analysis stratifying patients who met or did not meet the conventional indication for ICD showed a better discriminative ability of the ECG-AI index with 0.64 (95% CI, 0.53–0.75) of ROC-AUC in those who did not meet the ICD indication (n = 809) compared with those who met the ICD indication (ROC-AUC = 0.51 [95% CI, 0.38–0.65]).

### Competing Risk Analysis for Adjusting Non-SCD

The results of the survival analysis accounting for the competing risk of non-SCD (eTable 2 in the **Supplement**) show that the ECG-AI index (adjusted sub-distributional hazard ratio [sHR], 1.23; 95% CI, 1.04–1.49; p = 0.015), as well as the conventional guideline–directed indication for ICD (adjusted sHR, 1.98; 95% CI, 1.11–3.54; p = 0.02), were independently and significantly associated with the risk of SCD using the Fine-Gray models. In this competing analysis, our new model combined with the conventional indication for ICD and ECG-AI index showed good discriminative ability, with a 0.65 concordance index (95% CI, 0.62–0.69) using a bootstrapping technique (500 sets).

In subgroup analyses, the ECG-AI index was also independently associated with composite SCD events among each subset of patients in the Fine-Gray competing risk model (eFigure 5 in the **Supplement**). The cumulative incidence calculated via Aalen-Johansen estimates and further demonstrates this relationship (eFigure 6, using both the ECG-AI index (A) and the ECG-AI combined model with LVEF and NYHA class (B), in the **Supplement**). These results were similar to the time-dependent ROC-AUC without adjustment for the competing risk of non-SCD (eFigure 7 in the **Supplement**).

### Proportional Risk of SCD vs. Non-SCD using the ECG-Based AI Models

Figure 4 shows that the ECG-AI index alone and the ECG-AI combined model could discriminate between SCD and non-SCD across the low-, intermediate-, and high-risk patient groups. We observed an increase in the proportional risk of SCD vs. non-SCD as the ECG-AI index increased as follows: low risk, 16.7%; intermediate risk, 18.5%; high risk, 28.7% (p for trend = 0.023). A similar but sharper separation was seen in the ECG-AI combined model: low risk, 11.8%; intermediate risk, 15.7%; high risk, 36.1% (p for trend < 0.001).

## DISCUSSION

In the present study, the association between the ECG-AI index and SCD was evaluated in consecutive patients who required hospitalization for HF. Overall, the ECG-AI index, when added to the conventional guideline–directed indication for ICD based on LVEF and NYHA functional class, significantly improved the indices of discrimination and reclassification of SCD. We also observed an increased proportional risk estimation of SCD versus non-SCD. Importantly, similar findings were observed in subsets of patients with HF with a non-ischemic etiology and those with an LVEF > 35%.

The early AI model applied to the ECG data used neural network structures (i.e., multilayer perceptron) other than CNN or RNN. Improvements in computing and neural network technology have allowed the development of a deeper network pattern and, as a result, have enabled the handling of more complex data. For example, ECG-AI models using two-dimensional CNN reportedly predict age and sex and detect LV function and further latent atrial fibrillation from normal sinus rhythm ECG.^16,17,24^ More recently, we reported that an ECG-based AI model combining a one-dimensional CNN with RNN (i.e., LSTM) successfully identified patients with chest pain requiring urgent revascularization in an emergency setting.^22^ RNN can theoretically learn the time-series voltage data more precisely than CNN, as it explicitly deals with data ordering.^25^ Although some complex tasks may still require an RNN, the superiority of model performance using RNN over CNN is unclear; thus, consensus is lacking about the tasks that are suitable for RNN or CNN. In the present study, we attempted to establish an ECG-AI model using RNN combined with CNN to predict the incidence of SCD in patients with HF, and our results showed good performance beyond the conventional indication for ICD. Accurate risk prediction of SCD is essential in clinical practice, and these new approaches using AI algorithms may help clinicians provide a basis for decision-making to ensure the appropriate application of ICDs.

Previous studies reported that several ECG features (e.g., heart rate variability, T-wave alternance, early repolarization, late potential, and atrial fibrillation) are associated with SCD and can provide independent values beyond LV dysfunction.^26,27^ In fact, the Grad-CAM showed that our neural network model looked at the QRS wave and T wave, which seemed to focus on early repolarization, late potential, and T-wave alternance. Our ECG-based AI algorithm likely integrates specific features associated with SCD and can be accurately applied to a broader spectrum of patients with HF than classical predictive models. This is important as SCD represents a substantial burden for patients with HF and an LVEF > 35%, as evidenced by the fact that approximately 50% of SCD cases occur in the absence of severe LV dysfunction following myocardial infarction.^28^ Furthermore, a retrospective analysis of 714 patients with SCD found that only one-third exhibited sufficient LV dysfunction to meet the ICD criteria.^29^ Our ECG-based AI algorithm demonstrated a more refined risk stratification, in particular, an enrichment in the proportional risk estimation of SCD vs. non-SCD regardless of LVEF. As the prediction of SCD in these patient populations is considered highly difficult, we believe that our findings are highly encouraging. The number of patients with HF and an LVEF > 35% is increasing worldwide; therefore, we believe that our ECG-based AI algorithm will play a significant role in future research and clinical practice.

A known limitation of the conventional ICD indication also exists in non-ischemic patients. The DANISH trial (a study to assess the efficacy of ICDs in patients with non-ischemic systolic HF on mortality) reported that patients with HF of a non-ischemic etiology could not benefit from ICD as a primary prevention for SCD.^30^ However, a subgroup analysis of the DANISH trial indicated a mortality benefit from ICD implantation in younger patients, corresponding to a lower proportion of SCD relative to non-SCD with increasing age.^31^ In the present study, the ECG-AI index showed good discriminative ability, especially among patients with a non-ischemic etiology. Given the need to maximize the clinical benefit of ICDs, our ECG-based AI algorithm has the potential to aid in the identification of patients at high risk of SCD who are not currently captured by existing tools.

We also observed that the ECG-AI index alone and the ECG-AI combined model could discriminate between SCD and non-SCD across patients with different risk scores. As patients at increased risk of SCD are also likely to have a significantly higher risk of non-SCD mortality, those with a higher absolute risk of SCD are not always at a higher proportional risk. Importantly, previous reports demonstrated that the projected ICD benefit is relatively insensitive to absolute SCD risk but is highly sensitive to proportional risk.^10,32^ Furthermore, a recent analysis identified seven novel indicators associated with SCD; however, all were associated with non-SCD to at least the same extent and, hence, do not specifically predict SCD.^33^ These observations further highlight the potential impact of the ECG-AI index applied in our study. Overall, the ECG-AI index appears well poised to meet the requirements of SCD predictions, which are distinctly lacking from conventional standards.

### Strengths and Limitations

This study has some strengths, including the standardized assessment and adjudication of SCD by the central study committee of the WET-HF registry and the derivation and validation of the ECG-AI index using different hospitals with different patient backgrounds. This study also has several inherent limitations. First, the limited number of patients and clinical events resulted in a relatively low power to detect the incidence of fatal arrhythmic events, although our results are comparable to previously reported values demonstrating that ∼20% of patients with HF with a preserved LVEF succumbed to SCD over a 3-year follow-up period.^29^ In addition, we did not perform substantial statistical adjustments in the multivariable models, in which only two parameters, LVEF and NYHA class, were covariables. Other indicators, such as sex and body mass index, are reportedly useful for discriminating SCD from non-SCD,^10^ but they have not been universally confirmed as relevant tools for predicting SCD. We believe that our ECG-based AI algorithm adds substantial value to the current strategy for risk prediction of SCD with extremely low accuracy. Finally, a pitfall of these AI models is that unidentified biases or flaws can exist in the dataset, which can lead to misclassification. Further investigations are needed to validate our results in external cohorts with high-quality data inputs and ultimately compare AI-guided treatment with the standard treatment in a randomized controlled trial.

## CONCLUSIONS

The multifactorial nature of the ECG-AI index has allowed the creation of a more sensitive predictive model that may address the current shortcomings of capturing dynamic and proportional SCD risk in patients with HF. In this study, the AI-based assessment of ECG was tested as a new model for risk stratification of SCD in patients with HF and was found to be more discriminatory than conventional standards. Specifically, we observed improved prediction of SCD in patients with an LVEF of 35%–50% and a non-ischemic etiology, as well as discrimination between SCD and non-SCD. The ECG-AI algorithm may offer a powerful tool for improving clinical decision-making regarding preventive treatment, including the efficient use of ICD implants.

## Data Availability

All data produced in the present study are available upon reasonable request to the authors.

## ACKNOWLEDGMENT

Editorial support, in the form of medical writing, assembling tables, and creating high-resolution images based on authors’ detailed directions, collating author comments, copyediting, fact checking, and referencing, was provided by Editage, Cactus Communications.

## Data Availability Statement

The data underlying this article will be shared upon reasonable request to the corresponding author.

