## Supplemental Materials for "Electrocardiography-Based Prediction of Sudden Cardiac Death in Heart Failure Patients: Application of Artificial Intelligence"

**Supplementary Appendix**

**Authors:**

Yasuyuki Shiraishi,^1*^ Shinichi Goto,^2*^ Nozomi Niimi,^1^ Yoshinori Katsumata,^3^

Ayumi Goda,^4^ Makoto Takei,^5^ Mike Saji,^6^ Yosuke Nishihata,^7^ Motoaki Sano,^1^

Keiichi Fukuda,^1^ Takashi Kohno,^4^ Tsutomu Yoshikawa,^6^ and Shun Kohsaka^1^

* These authors contributed equally to this work

**Affiliations:**

^1^Department of Cardiology, Keio University School of Medicine, Tokyo, Japan

^2^One Brave Idea and Division of Cardiovascular Medicine, Department of Medicine, Brigham and Women’s Hospital, Boston, MA, USA

^3^Institute for Integrated Sports Medicine, Keio University School of Medicine, Tokyo, Japan

^4^Department of Cardiovascular Medicine, Kyorin University School of Medicine, Tokyo, Japan

^5^Department of Cardiology, Saiseikai Central Hospital, Tokyo, Japan

^6^Department of Cardiology, Sakakibara Heart Institute, Tokyo, Japan

^7^Department of Cardiology, St. Luke’s International Hospital, Tokyo, Japan

**Table of Contents**

**eMethod 1.** Design of the WET-HF registry.

**eMethod 2.** Statistical analysis.

**eTable 1**. Sensitivity analyses of the discriminative ability of the ECG-AI index, and the frequency of SCD and non-SCD by LVEF categories.

Abbreviations: ECG-AI: Electrocardiography-based artificial intelligence, ROC-AUC: Receiver operating characteristic area under the curve, SCD: Sudden cardiac death, LVEF: Left ventricular ejection fraction, CI: Confidence interval.

**eTable 2.** Subdistributional HRs of the ECG-AI index for predicting SCD events.

Subdistributional HRs are derived from the Fine-Gray models, accounting for the competing risk of non-SCD.

Abbreviations: HR: Hazard ratio, CI: Confidence interval, ECG-AI: Electrocardiogram-based artificial intelligence, SCD: Sudden cardiac death, ICD: Implantable cardioverter-defibrillator, LVEF: Left ventricular ejection fraction, NYHA: New York Heart Association.

**eFigure 1**. Patient allocation flow chart.

Abbreviations: WET-HF: West Tokyo Heart Failure, ECG: Electrocardiogram.

**eFigure 2**. Architecture of the neural network model.

Abbreviations: ECG: Electrocardiogram, 1D: One-dimensional, LSTM: Long short-term memory, SCD: Sudden cardiac death.

**eFigure 3**. Schematic illustration of the process of training and testing the neural network model.

The model was trained with data from the derivation cohort, and the performance of each model was calculated using data from the validation dataset at the end of each epoch. The final model was chosen as the model that performed best for 50 epochs in the validation cohort. The performance of the final model was calculated only once using data from the testing dataset.

**eFigure 4.** Visualization of the region of interest that our final neural network model interpreted based on Grad-CAM results.

**eFigure 5.** Forest plots of subdistributional HRs and 95% CI of the ECG-AI index for SCD by each subgroup.

**eFigure 6.** Cumulative incidence of SCDs and non-SCDs according to estimated risk by the ECG-AI models.

The cutoffs of SCD risk were defined by the Youden index of the logistic regression model for predicting 3-year SCD: (a) ECG-AI index, and (b) ECG-AI combined model with LVEF and NYHA class.

Abbreviations: SCD: Sudden cardiac death, ECG-AI: Electrocardiogram-based artificial intelligence, LVEF: Left ventricular ejection fraction, NYHA: New York Heart Association.

**eFigure 7.** Time-dependent ROC-AUC for predicting 3-year SCD events in the Cox proportional hazards models.

Abbreviations: ROC: Receiver operating characteristic area under the curve, SCD: Sudden cardiac death.

**eMethod 1**. Design of the WET-HF registry.

The West Tokyo Heart Failure Registry (WET-HF) was launched in January 2006 and consisted of six tertiary care hospitals as of December 2017. To ensure a robust assessment of the care and patient outcomes, baseline data and outcome measures were collected from the patients’ medical records, and the treating physicians were questioned by dedicated clinical research coordinators. Data were entered into an electronic data capturing system, which has a robust data query engine and system validations for data quality. Exclusive on-site auditing by the investigators (Y.S. and S.K.) ensured proper registration of each patient. The objective and detailed design was provided by the University Hospital Medical Information Network (UMIN000001171). The study protocol was approved by the ethical review committee of each center, and the study was conducted in accordance with the Declaration of Helsinki. According to the Ethical Guidelines for Medical and Health Research Involving Human Subjects and Personal Information Protection Law in Japan, informed consent was obtained from each participant prior to study initiation.

Patient demographics, medical history, laboratory and other tests (such as electrocardiogram and echocardiogram), medications, procedures, and clinical outcomes during hospitalization and after discharge with a minimum 2-year follow-up were recorded. Individual cardiologists made the clinical diagnosis of acute HF at each institution based on the Framingham criteria.^1^ In this cohort, NYHA functional class was evaluated at discharge by individual cardiologists at each institution and reviewed by the investigators (Y.S., S.K., T.K., Y.N., A.G., and T.Y.). LVEF on echocardiography was assessed using the modified Simpson’s method during the index hospitalization after stabilization of the HF signs and symptoms.

1. McKee PA, Castelli WP, McNamara PM, Kannel WB. The natural history of congestive heart failure: the Framingham study. *N Engl J Med*. 1971;285(26):1441-1446.

**eMethod 2.** Statistical analysis.

With respect to descriptive statistics, continuous variables are presented as median and interquartile range, while categorical variables are presented as frequency and percentage. For baseline characteristics, the three cohorts (i.e., derivation, validation, and test) were compared using the Kruskal–Wallis rank sum test for continuous variables and the chi-square test or Fisher’s exact test for categorical variables as appropriate.

| LVEF category | **Frequency** | | | **ECG-AI index performance** | |
| --- | --- | --- | --- | --- | --- |
|  | **Patient, number** | **SCD, number (%)** | **Non-SCD, number (%)** | **ROC-AUC** | **95% CI** |
| **LVEF cutoff: 45%** |  |  |  |  |  |
| ≤ 35% | 291 | 25 (8.6) | 49 (16.8) | 0.53 | 0.40–0.66 |
| 35%–45% | 215 | 13 (6.0) | 51 (23.7) | 0.61 | 0.40–0.81 |
| ≥ 45% | 571 | 21 (3.7) | 124 (21.7) | 0.63 | 0.50–0.76 |
| **LVEF cutoff: 50%** |  |  |  |  |  |
| ≤ 35% | 291 | 25 (8.6) | 49 (16.8) | 0.53 | 0.40–0.66 |
| 35%–50% | 329 | 21 (6.4) | 73 (22.2) | 0.69 | 0.55–0.83 |
| ≥ 50% | 457 | 13 (2.8) | 102 (22.3) | 0.53 | 0.38–0.69 |
| **LVEF cutoff: 55%** |  |  |  |  |  |
| ≤ 35% | 291 | 25 (8.6) | 49 (16.8) | 0.53 | 0.40–0.66 |
| 35%–55% | 436 | 26 (6.0) | 100 (22.9) | 0.65 | 0.52–0.78 |
| ≥ 55% | 350 | 8 (2.3) | 75 (21.4) | 0.51 | 0.34–0.68 |
| **LVEF cutoff: 60%** |  |  |  |  |  |
| ≤ 35% | 291 | 25 (8.6) | 49 (16.8) | 0.53 | 0.40–0.66 |
| 35%–60% | 572 | 31 (5.4) | 129 (22.6) | 0.66 | 0.55–0.77 |
| ≥ 60% | 214 | 3 (1.4) | 46 (21.5) | 0.58 | 0.38–0.77 |

**eTable 1.** Sensitivity analyses of the discriminative ability of the ECG-AI index and frequency of SCD and non-SCD by LVEF categories

Abbreviations: ECG-AI: Electrocardiography-based artificial intelligence, ROC-AUC: Receiver operating characteristic area under the curve, SCD: Sudden cardiac death, LVEF: Left ventricular ejection fraction, CI: Confidence interval.

**Table S2.** Subdistributional HRs of the ECG-AI index for predicting SCD events

| Characteristic | **Univariable** | | | **Multivariable** | | |
| --- | --- | --- | --- | --- | --- | --- |
|  | **sHR** | **95% CI** | **P-value** | **sHR** | **95% CI** | **P-value** |
| **Indication for ICD** |  |  |  |  |  |  |
| LVEF ≤ 35% and NYHA II–III | 2.25 | 1.30–3.89 | 0.004 | 1.98 | 1.11–3.54 | 0.020 |
| **ECG-AI index** |  |  |  |  |  |  |
| Z-score (standardized) | 1.31 | 1.12–1.53 | 0.001 | 1.23 | 1.04–1.49 | 0.015 |

Subdistributional HRs are derived from the Fine-Gray models, accounting for the competing risk of non-SCD.

Abbreviations: HR: Hazard ratio, CI: Confidence interval, ECG-AI: Electrocardiogram-based artificial intelligence, SCD: Sudden cardiac death, ICD: Implantable cardioverter-defibrillator, LVEF: Left ventricular ejection fraction, NYHA: New York Heart Association.

**Figure S1**. Patient allocation flow chart

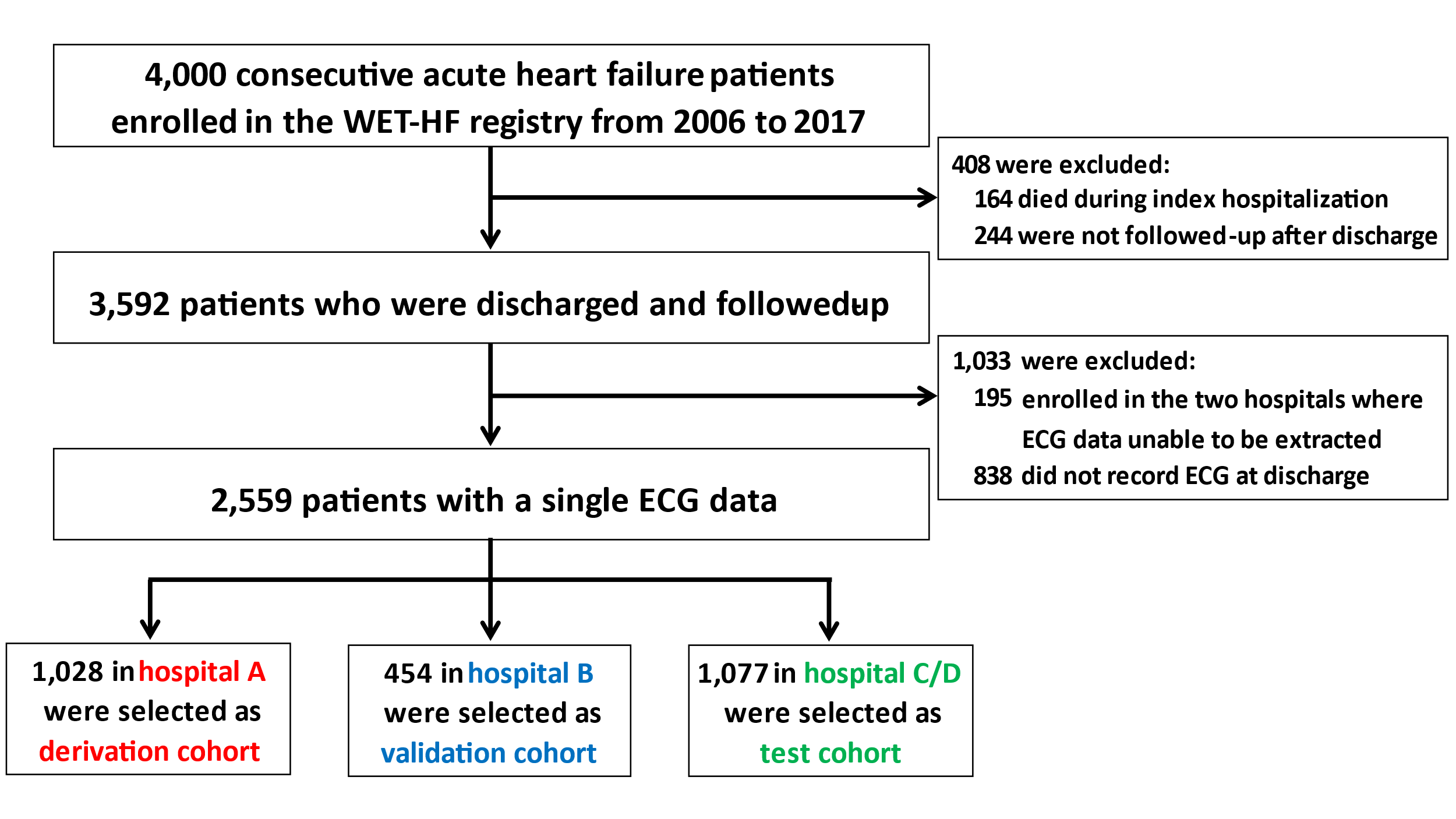

Abbreviations: WET-HF: West Tokyo Heart Failure, ECG: Electrocardiogram.

**Figure S2**. Architecture of the neural network model

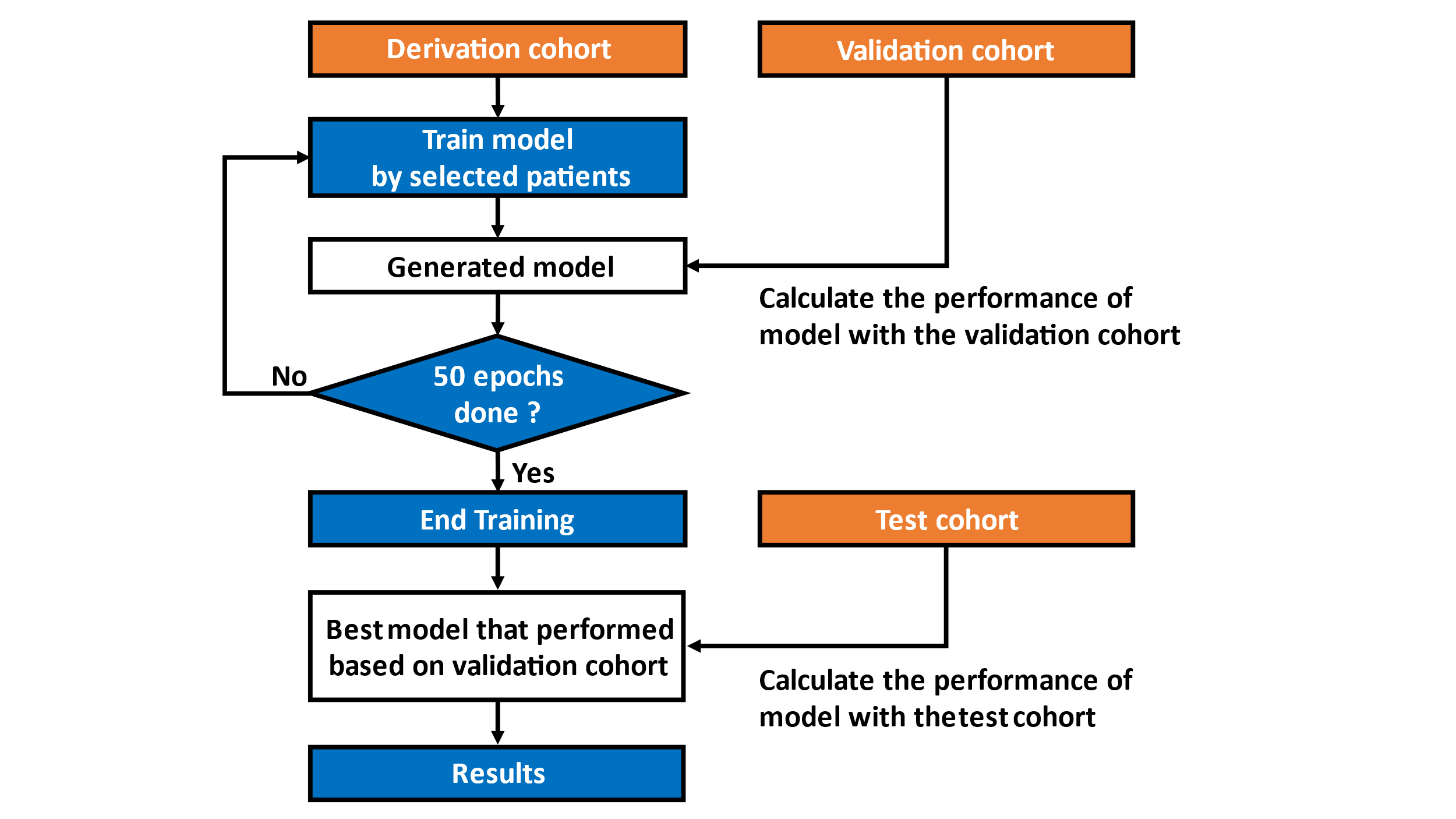

Abbreviations: ECG: Electrocardiogram, 1D: One-dimensional, LSTM: Long short-term memory, SCD: Sudden cardiac death.

**Figure S3.** Schematic illustration of the process of training and testing the neural network model

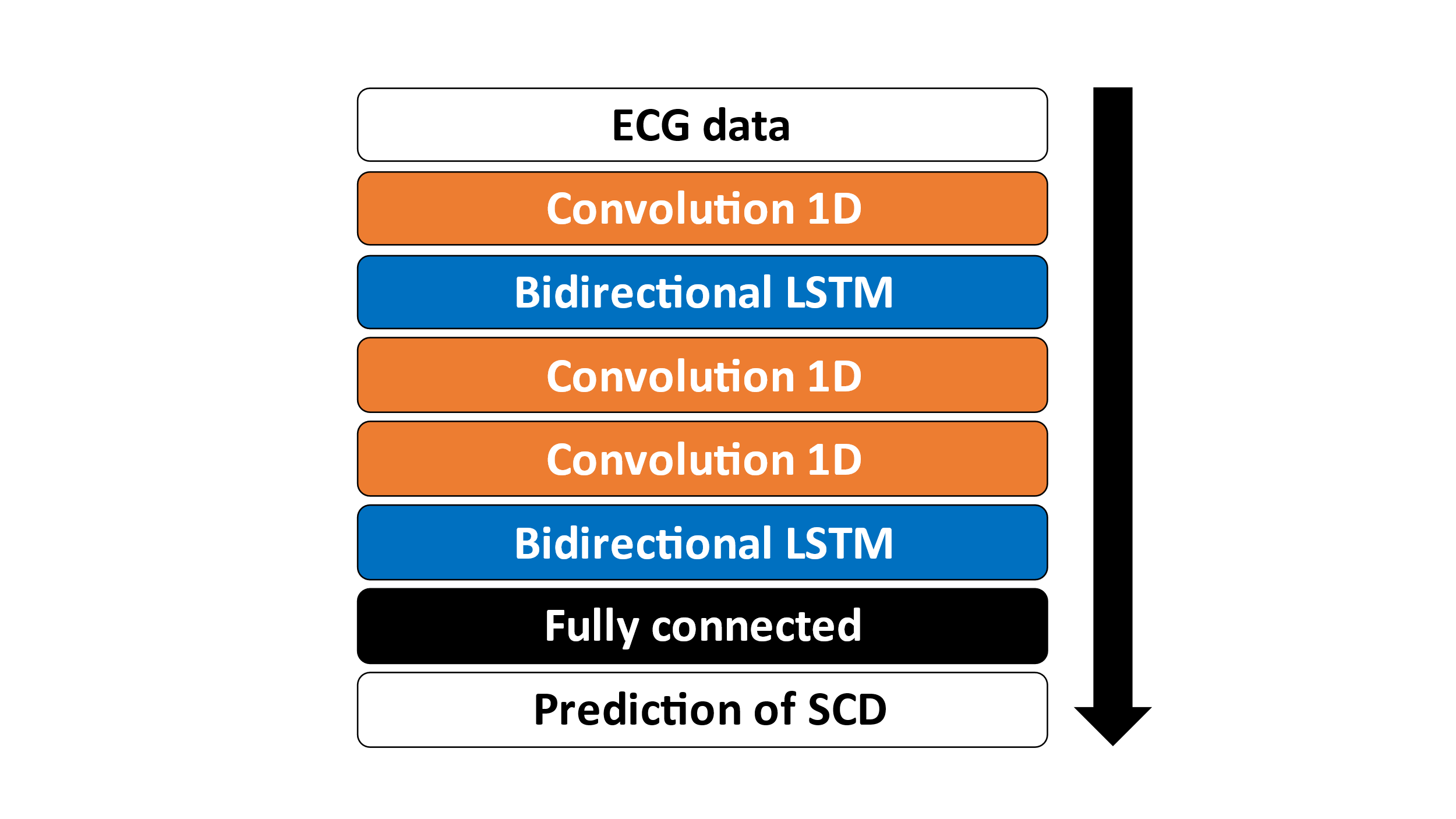

The model was trained with data from the derivation cohort, and the performance of each model was calculated using data from the validation dataset at the end of each epoch. The final model was chosen as the model that performed best for 50 epochs in the validation cohort. The performance of the final model was calculated only once using data from the testing dataset.

**Figure S4.** Visualization of the region of interest that our final neural network model interpreted based on Grad-CAM results.

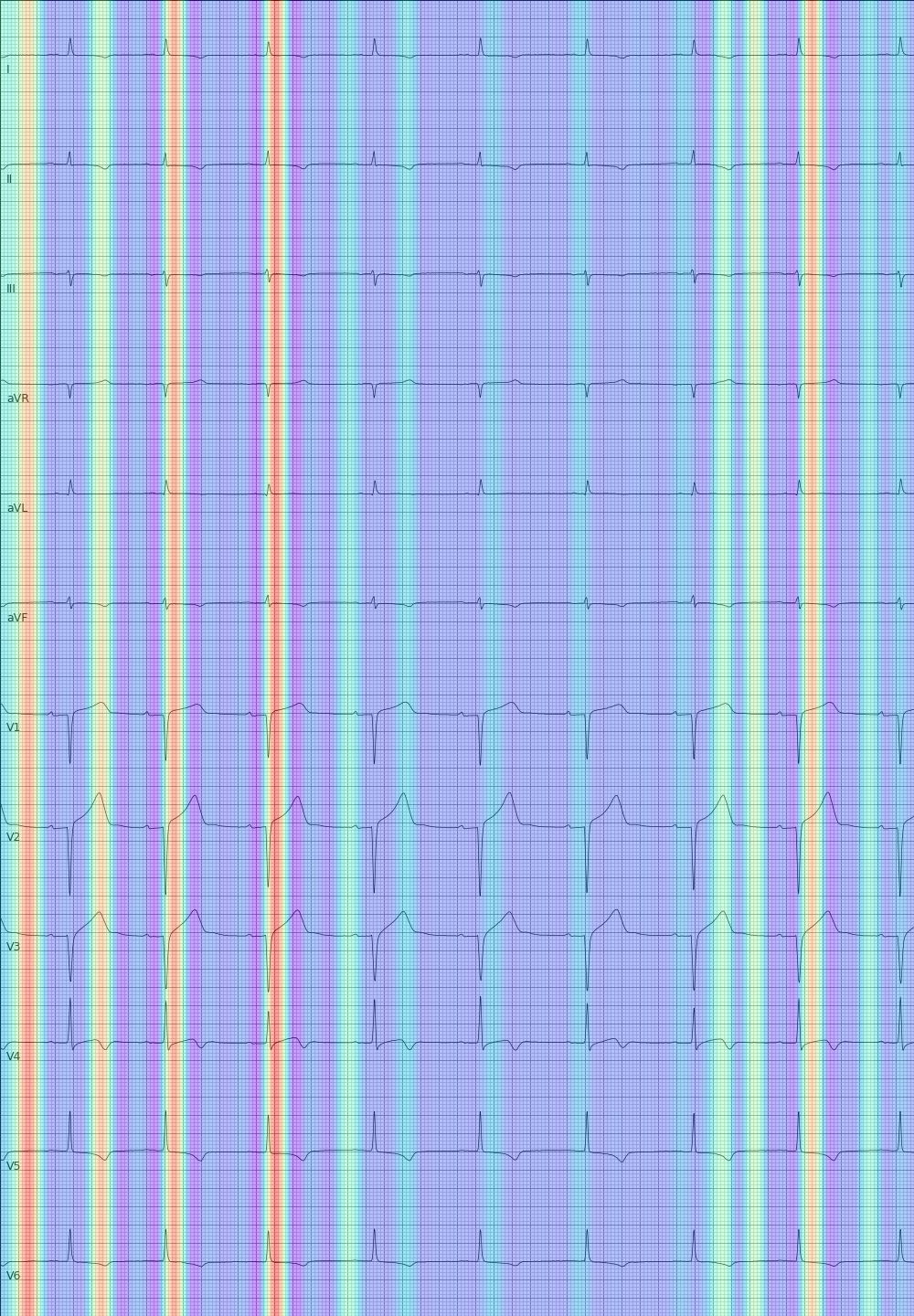

**Figure S5.** Forest plots of subdistributional HRs and 95% CI of the ECG-AI index for SCD by each subgroup

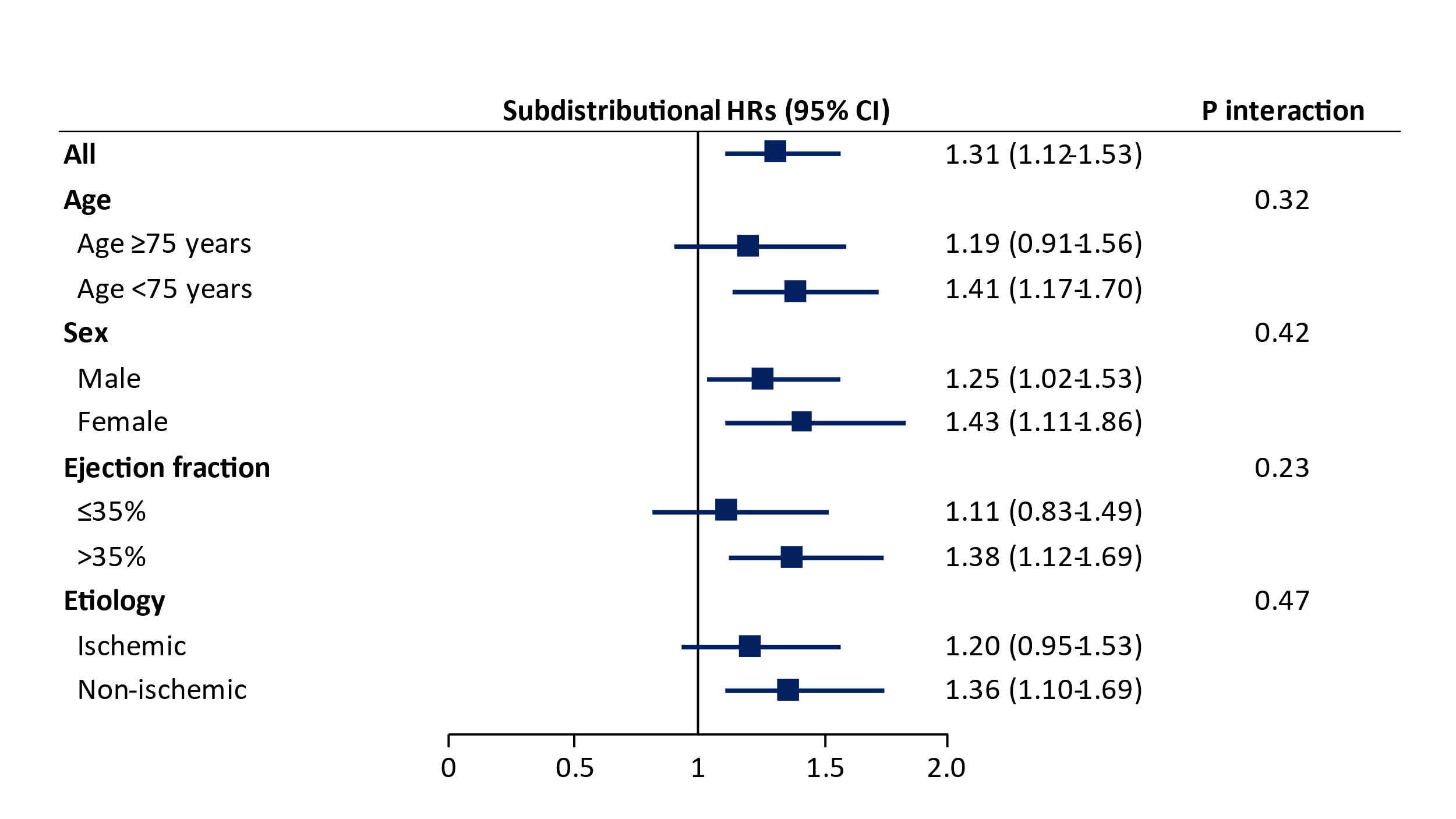

Abbreviations: HR: Hazard ratio, CI: Confidence interval, ECG-AI: Electrocardiogram-based artificial intelligence.

**Figure S6.** Cumulative incidence of SCDs and non-SCDs according to estimated risk by the ECG-AI models

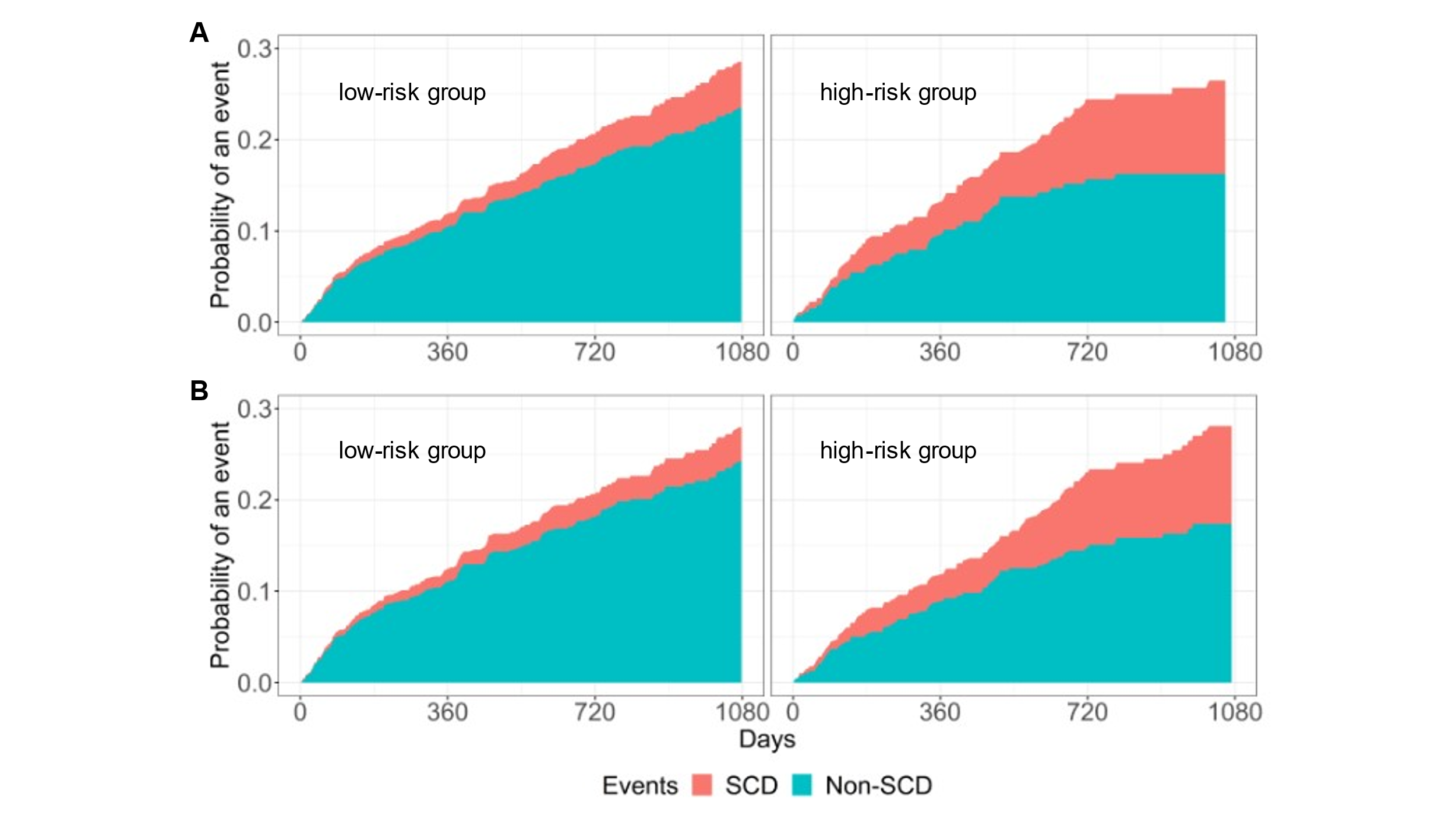

The cutoffs of SCD risk were defined by the Youden index of the logistic regression model for predicting 3-year SCD: (a) ECG-AI index, and (b) ECG-AI combined model with LVEF and NYHA class.

Abbreviations: SCD: Sudden cardiac death, ECG-AI: Electrocardiogram-based artificial intelligence, LVEF: Left ventricular ejection fraction, NYHA: New York Heart Association.

**Figure S7**. Time-dependent ROC-AUC for predicting 3-year SCD events in the Cox Proportional Hazards models

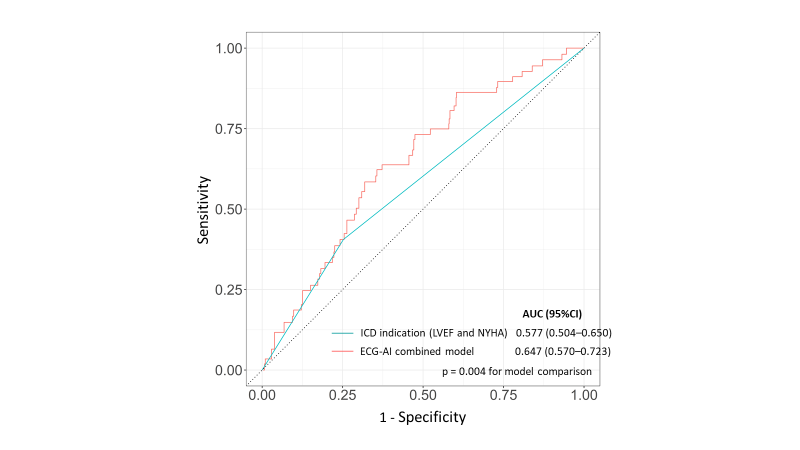

Abbreviations: ROC-AUC: Receiver operating characteristic area under the curve, SCD: Sudden cardiac death.
